# Optimal dose and safety of molnupiravir in patients with early SARS-CoV-2: a phase 1, dose-escalating, randomised controlled study

**DOI:** 10.1101/2021.05.03.21256309

**Authors:** Saye H Khoo, Richard FitzGerald, Thomas Fletcher, Sean Ewings, Thomas Jaki, Rebecca Lyon, Nichola Downs, Lauren Walker, Olana Tansley-Hancock, William Greenhalf, Christie Woods, Helen Reynolds, Ellice Marwood, Pavel Mozgunov, Emily Adams, Katie Bullock, Wayne Holman, Marcin D Bula, Jennifer L Gibney, Geoffrey Saunders, Andrea Corkhill, Colin Hale, Kerensa Thorne, Justin Chiong, Susannah Condie, Henry Pertinez, Wendy Painter, Emma Wrixon, Lucy Johnson, Sara Yeats, Kim Mallard, Mike Radford, Keira Fines, Victoria Shaw, Andrew Owen, David G Lalloo, Michael Jacobs, Gareth Griffiths

## Abstract

**Background:** AGILE is a phase Ib/IIa platform for rapidly evaluating COVID-19 treatments. In this trial (NCT04746183) we evaluated the safety and optimal dose of molnupiravir in participants with early symptomatic infection.

**Methods:** We undertook a dose-escalating, open-label, randomised-controlled (standard-of-care) Bayesian adaptive phase I trial at the Royal Liverpool and Broadgreen Clinical Research Facility. Participants (adult outpatients with PCR-confirmed SARS-CoV-2 infection within 5 days of symptom onset) were randomised 2:1 in groups of 6 participants to 300mg, 600mg and 800mg doses of molnupiravir orally, twice daily for 5 days or control. A dose was judged unsafe if the probability of 30% or greater dose-limiting toxicity (the primary outcome) over controls was higher than 25%. Secondary outcomes included safety, clinical progression, pharmacokinetics and virologic responses.

**Results:** Of 103 volunteers screened, 18 participants were enrolled between 17 July and 30 October 2020. Molnupiravir was well tolerated at 400, 600 or 800mg doses with no serious or severe adverse events. Overall, 4 of 4 (100%), 4 of 4 (100%) and 1 of 4 (25%) of the participants receiving 300, 600 and 800mg molnupiravir respectively, and 5 of 6 (83%) controls, had at least one adverse event, all of which were mild (≤grade 2). The probability of ≥30% excess toxicity over controls at 800mg was estimated at 0.9%.

**Conclusion:** Molnupiravir was safe and well tolerated; a dose of 800mg twice-daily for 5 days was recommended for Phase II evaluation.

## INTRODUCTION

In addition to life-saving therapies for COVID-19, there is an urgent need for effective antivirals in mild to moderate disease in order to reduce disease burden, prevent hospitalisation and death and potentially decrease transmission of SARS-CoV-2. AGILE is a randomised multi-arm, multi-dose, phase Ib/IIa platform in the UK using a seamless Bayesian adaptive design ^1^ to determine the safety, activity and optimal dose of multiple SARS-CoV-2 candidate therapeutics. Several candidates can be tested simultaneously (potentially sharing control group data) to increase efficiency.

We evaluated molnupiravir (EIDD-2801/MK-4482), for the treatment of COVID-19 in a seamless phase I/II trial. Molnupiravir is the prodrug of the ribonucleoside analogue 14 ß-d-N4-hydroxycytidine (NHC; EIDD-1931). Despite differences in model systems, activity of molnupiravir has consistently been demonstrated *in-vitro* and in animal models. In mice implanted with authentic human lung tissue, a prophylactic dose of 500 mg/kg given 12h prior to inoculation with SARS-CoV-2 and every 12h thereafter dramatically reduced viral plaque forming units at 2 days post inoculation.^2^ Furthermore, a twice daily 200mg/kg dose (but not 75mg/kg) was also able to reduce pulmonary viral RNA and improve lung histopathology in Syrian Golden Hamsters when initiated at the time of inoculation but with much lower efficacy if initiated ≥24h after infection.^3^ Finally, molnupiravir significantly reduced viral titres in the nasal swabs and turbinate 4 days after infection in ferrets when given at 5mg/kg twice daily initiated 12h after inoculation or 15mg/kg initiated 36h after inoculation,^4^ and was able to block transmission between ferrets. Current data warrant investigation of molnupiravir in human patients including studies to define the appropriate dose for a human SARS-CoV-2 antiviral indication.

Molnupiravir has been evaluated in healthy volunteers in single (50-1600mg) and multiple (50-800mg for 5.5 days) ascending oral doses, and was found to be well-tolerated ^5^. Preliminary data have also been presented from a study in patients with mild-to-moderate SARS CoV2 infection who received 200mg, 400mg or 800mg of molnupiravir twice daily for 5 days or placebo ^6^. Virus was cultured from nasopharyngeal swabs in only 42.9% of all PCR-positive patients at baseline and of these, culture-negativity was seen in all 47 evaluable subjects receiving molnupiravir (regardless of dose) versus 24% subjects allocated to placebo.

Here we report phase Ib results where we sought to determine the safety and tolerability of multiple ascending doses of molnupiravir in participants with symptomatic COVID-19 to recommend a dose for phase II. Secondary objectives included characterising adverse events (AEs), serious adverse events (SAEs), clinical outcomes (FLU-PRO, WHO Ordinal Scale, NEWS2 and mortality) as well as the pharmacokinetics of molnupiravir and its major metabolite EIDD-1931.

## METHODS

### Study design and Participants

This dose-escalation phase I study (NCT04746183) was designed as an open label, randomised, controlled Bayesian adaptive trial in adult early infection in the community, coordinated by the National Institute for Health Research (NIHR) Southampton Clinical Trials Unit with participants recruited into the NIHR Royal Liverpool and Broadgreen Clinical Research Facility (UK). Eligible participants were men and women aged ≥18 years with PCR-confirmed SARS-CoV-2 infection who were within 5 days of symptom onset, free of uncontrolled chronic conditions, and ambulant in the community with mild or moderate disease. Women of childbearing potential and men were required to use two effective methods of contraception, one of which should be highly effective, throughout the study and for 50 days and 100 days thereafter respectively. Any of the following criteria excluded participants from the study: pregnant or breast feeding women, stage 4 (severe) chronic kidney disease, clinically significant liver dysfunction, SpO2 <95% by oximetry or lung disease requiring supplementary oxygen, ALT and/or AST > 5 times upper limit of normal, platelets <50×10-9/L, experiencing any >= Grade 3 CTCAE v5 events, previously reported hepatitis C infection, known allergy to any study medication or having received any other experimental agents within 30 days of first dose of study drug. All participants provided written informed consent before enrolment. The study protocol was reviewed and approved by the UK Medicines and Healthcare product Regulatory Agency (MHRA) and West Midlands Edgbaston Research Ethics Committee.

### Randomisation and masking

Four sequential molnupiravir dosing tiers were defined *a priori* (300mg, 400mg, 600mg and 800mg BD for 5 days) with participants allocated using permuted blocks (block size 3, with no further stratification factors, generated by NIHR Southampton CTU statisticians) via MEDIDATA RAVE. Randomisation used a 2:1 allocation ratio so that within each cohort, 4 participants were randomly assigned to receive molnupiravir plus standard-of-care and 2 participants (controls) standard-of-care alone. The study was open label so both participant and treating clinician were aware of the allocated treatment.

### Procedures

Participants with laboratory-confirmed SARS-CoV-2 infection, or who had an illness compatible with COVID-19 (and who were subsequently confirmed to be positive) were screened against eligibility criteria, including presence and onset of symptoms within the previous 5 days. For safety reasons, in each cohort, the first participant randomised to molnupiravir (sentinel patient) was followed up until for 24 hours before any subsequent participants were randomised. All participants who received molnupiravir received drug after at least a two hour fasting period with a 4 hour period of observation after the first dose.

We utilised a Bayesian adaptive design to support decision making in this phase I study. Details are provided in Supplement S1. Briefly, a dose-toxicity model ^7^ was established which describes the relationship between dose-limiting toxicity at day 7 and treatment dose (control, 300mg, 400mg, 600mg and 800mg BD) and updated following completion of each dosing tier – see figure 2. For each cohort, the Safety Review Committee (SRC) reviewed all available safety data including at least 7 days data for each participant in the cohort, and all accrued information on previous cohorts (up to a maximum follow-up of 28 days). This included AE data, vital signs data, ECG data and clinical laboratory evaluations, as well as any emerging data from other studies. Following SRC review, recommendations could be to de-escalate, escalate, remain at the same dose, or continue to phase II. A dose was deemed to be unsafe if there was a ≥25% chance that treatment was associated with a >30% risk of dose-limiting toxicities at day 7. The model recommended the next dose-level according to which level is the most likely to correspond to an increase of a 15-25% in the dose limiting toxicity rate over control. However, the SRC made the ultimate decision whether to accept that the current dose was safe and to dose escalate and could decide to skip a dose if it did not more than double and was deemed safe by the Bayesian model. Once the dose escalation Phase I was complete, the independent Data Monitoring and Ethics Committee reviewed data from the final SRC, along with their recommendations on the recommended phase II dose, to ratify the Recommend Phase 2 Dose.

### Outcomes

The primary outcome was dose limiting toxicity (DLT) using CTCAE version 5 (grades 3 and above) measured over 7 days and CTCAE grading related to platelets and/or lymphocytes, assessed in all participants, who were randomly assigned and received at least one dose of molnupiravir (unless randomised to control). Secondary outcomes: for safety included AEs, SAEs, physical findings, vital signs and laboratory parameters; for pharmacokinetics included concentrations of molnupiravir and EIDD-1931 in plasma; for clinical included Patient Reported Outcome Measures (FLU-PRO), WHO COVID-19 Ordinal Scale (at days 15,29), NEWS2 (assessed during clinic days 15, 29), mortality (days 15, 29) and time from randomisation to death (up to day 29).

### Pharmacokinetic sampling

Plasma was sampled at Day 1 and 5 to measure the concentrations of molnupiravir and its major active metabolite EIDD-1931. On each sampling day, 2mL of venous blood was collected pre-dose, at 30 mins, and at 1-, 2- and 4-hours post-dose. All samples were rapidly cooled on wet ice and centrifuged (2000g for 10min) within 30 min of sample collection. Within 10 minutes of completing centrifugation, 150mL of plasma was mixed with 450uL of acetonitrile, vortexed and transferred to a -80°C freezer prior to onward shipping for pharmacokinetic analyses. Drug concentrations were measured using a validated LC-MS/MS assay at Covance Clinical Laboratories, Leeds, UK.

Concentrations of EIDD-1931 in plasma on day 1 and day 5 were described using summary statistics (geometric mean (90% CI), mean, standard deviation, median and range) for each time point.

Key pharmacokinetic (PK) parameters such as area under the concentration-time curve 0-4 h (AUC_0-4_), maximum concentration (Cmax) and time to maximum concentration (Tmax) were determined by non-compartmental modelling methods (WinNonlin, Phoenix, v. 8.3, Pharsight, Mountain View, CA, USA) on day 1 and day 5 for each dose and summarised descriptively. Accumulation ratios to day 5 were calculated for EIDD-1931 AUC_0-4_ and Cmax.

### Statistical analysis

All analyses are reported according to CONSORT 2010 and ICH E9 guidelines on Statistical Principles in Clinical Trials. All enrolled participants were included in both the evaluable population and the safety population for analysis.

The primary endpoint of DLTs up to 7 days post first dose were modelled using a Bayesian dose-toxicity model based on Mozgunov et al.^7^ The relationship between dose and toxicity was modelled using a two-parameter logistic model, where information can be shared across doses; in particular, the DLT rate in controls informs estimates for the active doses. The prior distributions for this model were calibrated to maximise the proportion of correct selection under a range of dose-toxicity scenarios where each dose considered in the study was the optimum one. The toxicity risk in controls was *a priori* assumed to be 10%. Further details are given in Supplement S1.

The dose-toxicity model was updated after every cohort of participants, and the final model is presented as estimated DLT rates for each dose, alongside equal-tail 95% credible intervals. For active doses, we also present estimated additional toxicity above controls, the probability that the DLT rate falls within 15-25% additional toxicity over controls (a pre-determined acceptable target range for toxicity) and the probability of at least 30% additional toxicity over controls (deemed as unacceptably toxic). This is supported by the same information for up to day 29.

Baseline demographics are summarised within each dose (and controls) using descriptive statistics. Clinical endpoints are similarly summarised at days 15 and 29. The sample size was flexible, based on the need for the study to adapt to accruing safety data. Simulations to assess model operating characteristics and to calibrate prior assumed four doses (plus controls), with cohorts of size six capped at a total of 30 participants. Statistical analysis was undertaken in SAS version 9·4, STATA version 16 and R version 3·6 ·0.

## RESULTS

Of 103 potential participants (Figure 1) who attended for screening, 58 were excluded (31 had no signs or symptoms of COVID-19, 12 tested negative for SARS-CoV-2 by PCR, 7 had signs or symptoms that began after 5 days of planned first dose, 2 had an uncontrolled comorbidity, 3 did not meet contraceptive requirements, 1 did not meet the age range, 1 did not meet the mild/moderate disease criterion, 22 declined, 5 were screened between dosing cohorts and 1 was unknown). Eligible individuals were randomly assigned within three sequential dose cohorts (300mg, 600mg & 800mg) of 6 participants each (i.e. a total of 18 participants within the phase 1) and dosed in the period between 17 July 2020, and 30 October 2020. The baseline characteristics of participants were similar across all groups (Table 1) with an overall median age of 56, 72% (13/18) female and 33% (6/18) having a WHO COVID ordinal score of 1 (ambulatory mild disease). The median number of days (range) from symptom onset to randomisation and treatment by the 18 participants was 4 (range 1-5).

**Figure 1:**
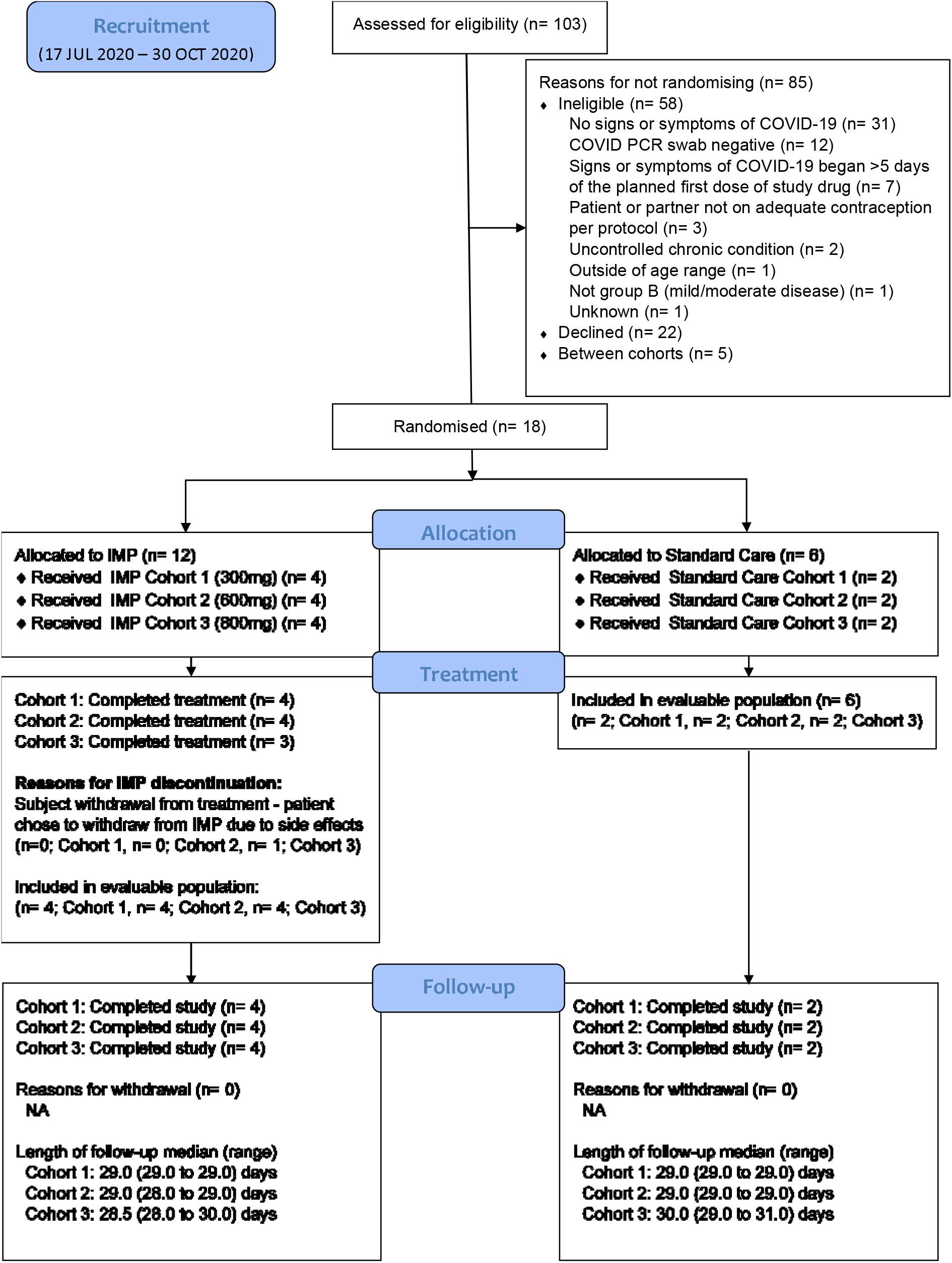
CONSORT diagram

**Figure 2:**
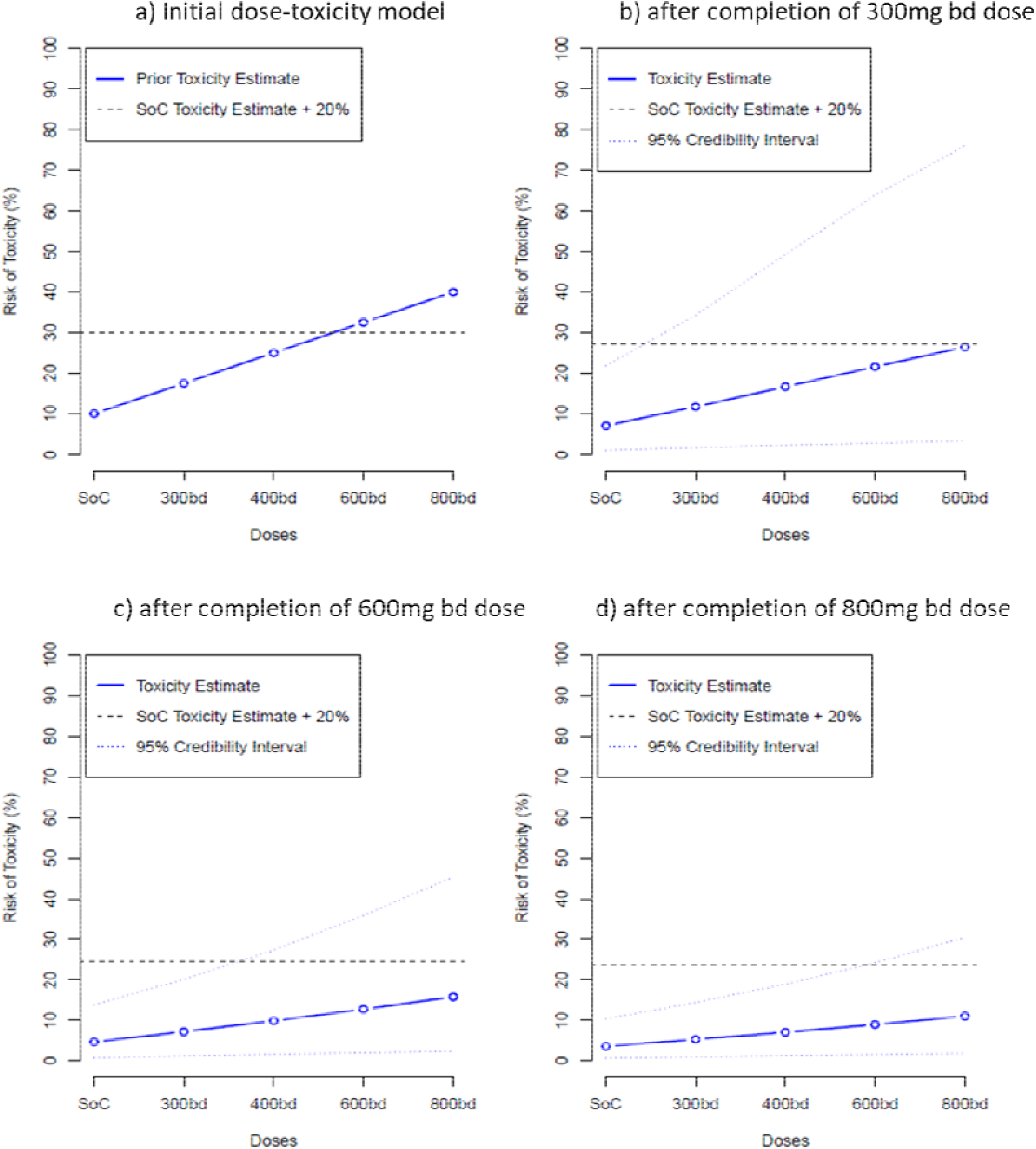
Primary endpoint - dose toxicity plot up to day 7 (evaluable population)

**TABLE 1:** Baseline characteristics.

|  | MOLNUPIR<br>AVIR<br>(300mg)<br>(n=4) | MOLNUPIR<br>AVIR<br>(600mg)<br>(n=4) | MOLNUPIR<br>AVIR<br>(800mg)<br>(n=4) | Standard<br>Care<br>Total<br>(n=6) | Total<br>(n=18) |
| --- | --- | --- | --- | --- | --- |
| **Age at consent (years)** |  |  |  |  |  |
| n | 4 | 4 | 4 | 6 | 18 |
| Median | 56.0 | 43.0 | 39.0 | 59.0 | 56.0 |
| Range | 51.0 to 80.0 | 22.0 to 60.0 | 25.0 to 63.0 | 22.0 to 63.0 | 22.0 to 80.0 |
| **Gender – n (%)** |  |  |  |  |  |
| Male | 1 (25.0%) | 2 (50.0%) | 0 (0.0%) | 2 (33.3%) | 5 (27.8%) |
| Female | 3 (75.0%) | 2 (50.0%) | 4 (100%) | 4 (66.7%) | 13 (72.2%) |
| **Ethnicity** |  |  |  |  |  |
| White - English / Welsh /<br>Scottish / Northern Irish | 4 (100%) | 4 (100%) | 4 (100%) | 6 (100%) | 18 (100%) |
| **BMI** |  |  |  |  |  |
| n | 4 | 4 | 4 | 6 | 18 |
| Median | 28.1 | 33.9 | 21.0 | 31.3 | 29.5 |
| Range | 25.6 to 32.7 | 30.0 to 51.1 | 20.4 to 34.0 | 27.2 to 36.2 | 20.4 to 51.1 |
| **WHO Score (day 1) – n (%)** |  |  |  |  |  |
| 1. Ambulatory mild disease,<br>asymptomatic; viral RNA<br>detected | 2 (50.0%) | 0 (0.0%) | 1 (25.0%) | 3 (50.0%) | 6 (33.3%) |
| 2. Ambulatory mild disease,<br>symptomatic; independent | 1 (25.0%) | 4 (100%) | 3 (75.0%) | 3 (50.0%) | 11 (61.1%) |

|  | MOLNUPIR | MOLNUPIR | MOLNUPIR | Standard |  |
| --- | --- | --- | --- | --- | --- |
|  | AVIR | AVIR | AVIR | Care | Total |
|  | (300mg) | (600mg) | (800mg) | Total | (n=18) |
|  | (n=4) | (n=4) | (n=4) | (n=6) |  |
| 3. Ambulatory mild disease,<br>symptomatic; assistance needed | 1 (25.0%) | 0 (0.0%) | 0 (0.0%) | 0 (0.0%) | 1 (5.6%) |
| **WHO Score (day 1)** |  |  |  |  |  |
| n | 4 | 4 | 4 | 6 | 18 |
| Median | 1.5 | 2 | 2 | 1.5 | 2 |
| Range | 1 to 3 | 2 to 2 | 1 to 2 | 1 to 2 | 1 to 3 |
| **NEWS2 Score (day 1)** |  |  |  |  |  |
| n | 4 | 4 | 4 | 6 | 18 |
| Median | 0 | 0 | 0.5 | 0 | 0 |
| Range | 0 to 1 | 0 to 0 | 0 to 1 | 0 to 0 | 0 to 1 |
| **O<sub>2</sub> Saturation % (day 1)** |  |  |  |  |  |
| n | 4 | 4 | 4 | 6 | 18 |
| Median | 97.5 | 96.5 | 99.0 | 98.0 | 97.5 |
| Range | 95.0 to 98.0 | 96.0 to 99.0 | 95.0 to 100.0 | 96.0 to 100.0 | 95.0 to 100 |
| **FLU-PRO total (day 1)** |  |  |  |  |  |
| n | 3 | 4 | 4 | 6 | 17 |
| Median | 0.7 | 0.8 | 1.0 | 0.6 |  |
| Range | 0.4 to 1.3 | 0.8 to 1.4 | 0.5 to 1.6 | 0.3 to 1.6 | 0.3 to 1.6 |
| Missing from eCRF – n (%) | 1 (25.0%) | 0 (0.0%) | 0 (0.0%) | 0 (0.0%) | 1 (5.6%) |
| **Time from symptom onset to**<br>**randomisation (days)<sup>2</sup>** |  |  |  |  |  |
| n | 4 | 4 | 4 | 6 | 18 |
|  | AVIR | AVIR | AVIR | Care | Total |
|  | (300mg) | (600mg) | (800mg) | Total | (n=18) |
|  | (n=4) | (n=4) | (n=4) | (n=6) |  |
| Median | 4.0 | 4.0 | 3.5 | 4.0 | 4.0 |
| Range | 3.0 to 4.0 | 4.0 to 4.0 | 2.0 to 4.0 | 1.0 to 5.0 | 1.0 to 5.0 |

**NOTE:** Percentages are based on the number of patients in the study arm
<sup>1</sup>Reason for missing FLU-PRO:
<sup>2</sup>**NOTE:** Date of randomisation is the same as date of first dose for all patients randomised to molnupiravir

All molnupiravir participants received at least 1 dose with 3/4 (75%), 4/4 (100%) and 3/4 (75%) completing the full treatment in the 300, 600 and 800mg cohort respectively. One participant on 300mg BD only took 1 of 2 intended tablets for 2 of their treatment doses and one participant on 800mg BD took only two doses on day one, withdrawing from treatment for personal reasons. The median number of molnupiravir doses received (range) was 10 (8-10), 10 (10-10), 10 (2-10) and median number of days on molnupiravir treatment (and range) was 5.5 (5-6), 5 (5-5), 5 (1-5) for the 300mg, 600mg & 800mg cohorts respectively.

### Primary analysis

No participants in any cohort experienced a DLT or a grade 3 or above change in lymphocytes or platelets (for those with a normal baseline value) or a 2 or more grade increment in lymphocytes or platelets (for those with grade 2 or 3 at baseline). Following review by the Safety Review Committee (SRC), dose cohort escalation went from 300mg to 600mg (skipping 400mg) and then from 600mg to 800mg. Bayesian model DLT point estimates, 95% credible interval, and the target toxicity level of 20% over the controls are shown in Figure 2. For data up to day 7, the maximum dose (800mg) had an estimated DLT rate of 11·0% (equal-tail 95% credible interval of 1·8 to 30·4%), with estimated 7·4% additional toxicity over controls and a probability of additional toxicity ≥30% over controls of 0.9%. As there were no DLTs recorded up to day 28, the results for day 7 are the same for day 28 and so are not repeated. These data support 800mg BD as the recommended phase II dose.

### Analysis of secondary endpoints

Adverse events were evenly distributed among the dose cohorts including controls. Overall, 4 of 4 (100%), 4 of 4 (100%) and 1 of 4 (25%) of the participants receiving 300, 600 and 800mg of molnupiravir, and 5 of 6 (83%) controls, had at least one adverse event, all of which were mild (≤ grade 2). Molnupiravir was generally well-tolerated compared with controls, and Table 2 describes the frequencies of events across the groups by system organ class and CTCAE term. No serious adverse events were reported. The most common symptoms were gastrointestinal (diarhoea, nausea), respiratory (cough), central nervous system (loss of smell or taste) and flu-like symptoms.

**TABLE 2:** Overall toxicity Summary by CTCAE version 5 term - Safety population.

| Characteristic | MOLNUPIRAVIR | MOLNUPIRAVIR | MOLNUPIRAVIR | Standard<br>Care |
| --- | --- | --- | --- | --- |
|  | (300mg)<br>(n=4) | (600mg)<br>(n=4) | (800mg)<br>(n=4) | Total<br>(n=6) |
| Number of patients that experienced at least one AE – n (%) * | 4 (100%) | 4 (100%) | 1 (25%) | 5 (83.3%) |
| Summary of AEs – n (%) |  |  |  |  |
| **Cardiac disorders** | **0 (0.0%)** | **0 (0.0%)** | **1 (25.0%)** | **0 (0.0%)** |
| Palpitations | 0 (0.0%) | 0 (0.0%) | 1 (25.0%) | 0 (0.0%) |
| **Ear and labyrinth disorders** | **0 (0.0%)** | **0 (0.0%)** | **0 (0.0%)** | **1 (16.7%)** |
| Tinnitus | 0 (0.0%) | 0 (0.0%) | 0 (0.0%) | 1 (16.7%) |
| **Eye disorders** | **0 (0.0%)** | **1 (25.0%)** | **0 (0.0%)** | **0 (0.0%)** |
| Blurred Vision | 0 (0.0%) | 1 (25.0%) | 0 (0.0%) | 0 (0.0%) |
| **Gastrointestinal disorders** | **2 (50.0%)** | **3 (75.0%)** | **0 (0.0%)** | **2 (33.3%)** |
| Abdominal Pain | 1 (25.0%) | 0 (0.0%) | 0 (0.0%) | 0 (0.0%) |
| Diarrhea | 2 (50.0%) | 1 (25.0%) | 0 (0.0%) | 1 (16.7%) |
| Dyspepsia | 0 (0.0%) | 1 (25.0%) | 0 (0.0%) | 0 (0.0%) |
| Nausea | 1 (25.0%) | 2 (50.0%) | 0 (0.0%) | 1 (16.7%) |
| Oral Dysesthesia | 1 (25.0%) | 0 (0.0%) | 0 (0.0%) | 0 (0.0%) |
| Vomiting | 1 (25.0%) | 0 (0.0%) | 0 (0.0%) | 0 (0.0%) |
| **General disorders and administration site conditions** | **1 (25.0%)** | **0 (0.0%)** | **0 (0.0%)** | **2 (33.3%)** |
| Fatigue | 1 (25.0%) | 0 (0.0%) | 0 (0.0%) | 0 (0.0%) |
| Flu Like Symptoms | 0 (0.0%) | 0 (0.0%) | 0 (0.0%) | 2 (33.3%) |
| Non-Cardiac Chest Pain | 0 (0.0%) | 0 (0.0%) | 0 (0.0%) | 1 (16.7%) |
|  | (300mg)<br>(n=4) | (600mg)<br>(n=4) | (800mg)<br>(n=4) | Total<br>(n=6) |
| **Infections and infestations** | **1 (25.0%)** | **2 (50.0%)** | **0 (0.0%)** | **0 (0.0%)** |
| Herpes Simplex Reactivation | 1 (25.0%) | 0 (0.0%) | 0 (0.0%) | 0 (0.0%) |
| Infections And Infestations - Other,<br>Specify<sup>1</sup> | 0 (0.0%) | 1 (25.0%) | 0 (0.0%) | 0 (0.0%) |
| Thrush | 0 (0.0%) | 1 (25.0%) | 0 (0.0%) | 0 (0.0%) |
| **Investigations** | **0 (0.0%)** | **1 (25.0%)** | **0 (0.0%)** | **0 (0.0%)** |
| Alanine Aminotransferase Increased | 0 (0.0%) | 1 (25.0%) | 0 (0.0%) | 0 (0.0%) |
| GGT Increased | 0 (0.0%) | 1 (25.0%) | 0 (0.0%) | 0 (0.0%) |
| **Musculoskeletal and connective**<br>**tissue disorders** | **1 (25.0%)** | **0 (0.0%)** | **0 (0.0%)** | **1 (16.7%)** |
| Chest Wall Pain | 1 (25.0%) | 0 (0.0%) | 0 (0.0%) | 0 (0.0%) |
| Myalgia | 0 (0.0%) | 0 (0.0%) | 0 (0.0%) | 1 (16.7%) |
| **Nervous system disorders** | **2 (50.0%)** | **1 (25.0%)** | **0 (0.0%)** | **2 (33.3%)** |
| Anosmia | 1 (25.0%) | 0 (0.0%) | 0 (0.0%) | 0 (0.0%) |
| Dizziness | 1 (25.0%) | 0 (0.0%) | 0 (0.0%) | 1 (16.7%) |
| Dysgeusia | 1 (25.0%) | 0 (0.0%) | 0 (0.0%) | 1 (16.7%) |
| Headache | 0 (0.0%) | 1 (25.0%) | 0 (0.0%) | 2 (33.3%) |
| **Psychiatric disorders** | **0 (0.0%)** | **0 (0.0%)** | **1 (25.0%)** | **0 (0.0%)** |
| Anxiety | 0 (0.0%) | 0 (0.0%) | 1 (25.0%) | 0 (0.0%) |
| **Renal and urinary disorders** | **1 (25.0%)** | **0 (0.0%)** | **0 (0.0%)** | **1 (16.7%)** |
| Chronic Kidney Disease | 0 (0.0%) | 0 (0.0%) | 0 (0.0%) | 1 (16.7%) |
| Urine Discoloration | 1 (25.0%) | 0 (0.0%) | 0 (0.0%) | 0 (0.0%) |

| Characteristic | MOLNUPIRAVIR | MOLNUPIRAVIR | MOLNUPIRAVIR | Standard |
| --- | --- | --- | --- | --- |
|  | (300mg) | (600mg) | (800mg) | Care |
|  | (n=4) | (n=4) | (n=4) | Total |
|  |  |  |  | (n=6) |
| **Respiratory, thoracic and mediastinal disorders** | **1 (25.0%)** | **1 (25.0%)** | **0 (0.0%)** | **1 (16.7%)** |
| Cough | 1 (25.0%) | 0 (0.0%) | 0 (0.0%) | 1 (16.7%) |
| Hoarseness | 0 (0.0%) | 1 (25.0%) | 0 (0.0%) | 0 (0.0%) |
| Rhinorrhea | 1 (25.0%) | 0 (0.0%) | 0 (0.0%) | 0 (0.0%) |
| Sore Throat | 0 (0.0%) | 1 (25.0%) | 0 (0.0%) | 0 (0.0%) |
| **Unclassified** | **1 (25.0%)** | **1 (25.0%)** | **1 (25.0%)** | **0 (0.0%)** |
| Other - Bilateral Thigh Pain | 1 (25.0%) | 0 (0.0%) | 0 (0.0%) | 0 (0.0%) |
| Other - Loose Stools | 0 (0.0%) | 0 (0.0%) | 1 (25.0%) | 0 (0.0%) |
| Other - Worsening Fatigue | 0 (0.0%) | 1 (25.0%) | 0 (0.0%) | 0 (0.0%) |

Note: Percentages are based on the number of patients in the study arm. CTCAE v5.0 terms are used to classify AEs.
<sup>1</sup>This AE reported in Other specify free text field as "Chest infection".
\* NB: All AEs were either Grade 1 or 2

At day 15 all participants had a WHO ordinal scale of 1 or 2, with a median score (range) of 1·5 (1-2), 1·5 (1-2), 2 (2-2) and 1·5 (1-2) for molnupiravir 300mg, 600mg and 800mg and controls respectively. At day 15 the molnupiravir 300mg, 600mg and controls had a median NEWS2 Score of 0 (range 0-0), with molnupiravir 800mg a median score of 1 (range 0-1). Median O2 saturation (range) was 97 (97-100), 97 (96-99), 99.5 (97-100), 97 (96-99) for 300, 600 and 800mg molnupiravir and controls respectively with median FLU-PRO totals 0·4 (0·2-10), 0·2 (0·1-0·6), 0·1 (0-0·3) and 0·2 (0-0·5) respectively (further details with comparable day 29 endpoints are provided in Supplement 2).

### Pharmacokinetics

The prodrug molnupiravir was generally not detectable, or detected at low concentrations only at early timepoints (0·5, 1 hour post-dose), at all 3 doses (Table 3). Plasma concentrations of the nucleoside metabolite EIDD-1931 were detectable, and showed no accumulation between day 1 and day 5. At day 5, geometric mean NHC exposures (%CV) over the first 4 hours of dosing (AUC_0-4_) in the 300mg (N=4), 600mg (N=4) and 800mg (N=3) dose were 3470 (42·4), 3880 (56·3) and 7880 (39·0) ng.h/mL, with corresponding peak (Cmax) concentrations of 1620 (51·0), 1820 (84·6) and 4180 (28·1) ng/mL. Time to peak plasma concentration was 0·5 – 2·0 hours.

**TABLE 3:**
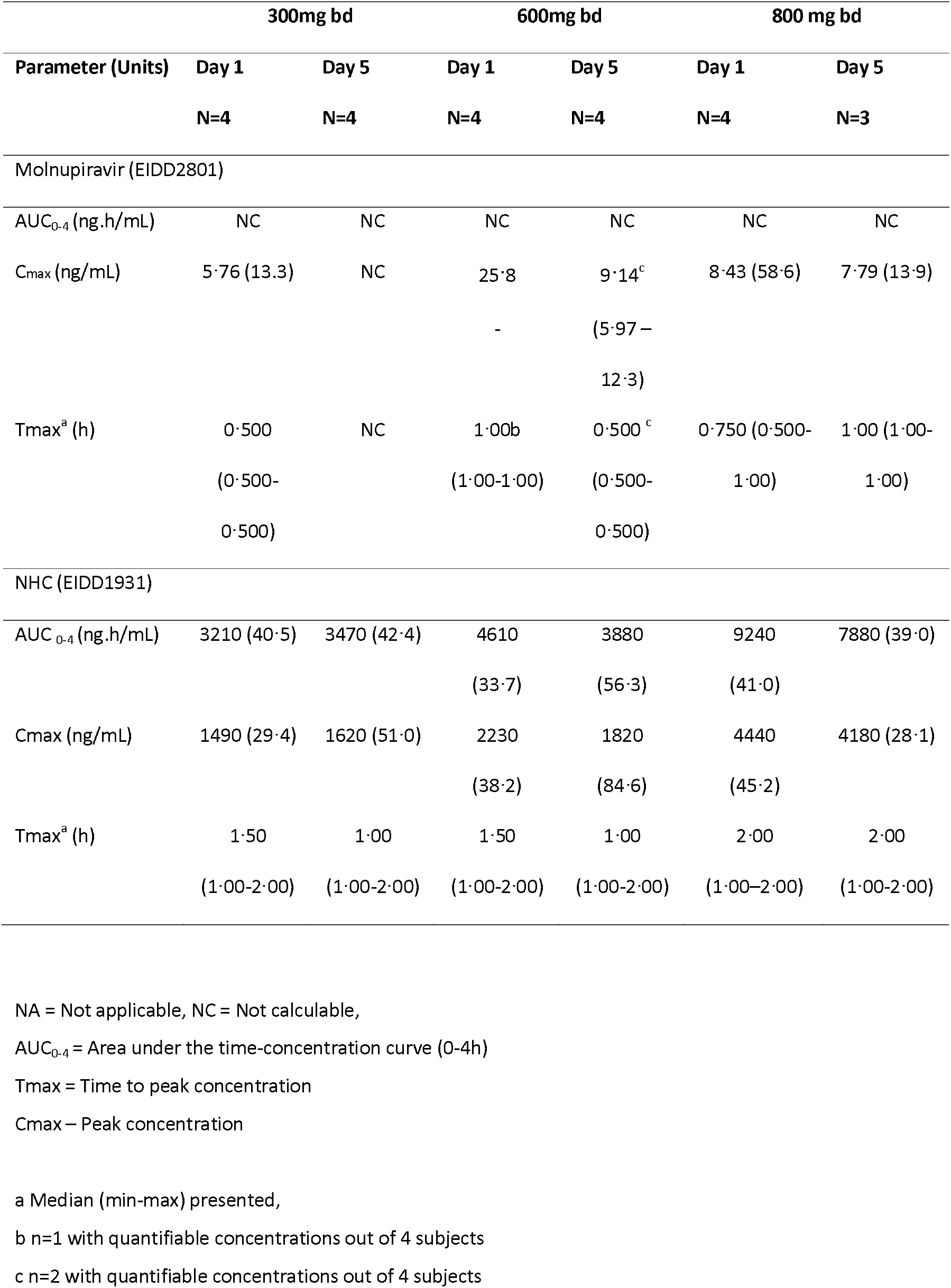
Geometric mean (%CV) Pharmacokinetic parameters of EIDD-2801 following single and multiple dose administration of EIDD-2801.

|  | 300mg bd |  | 600mg bd |  | 800 mg bd |  |
| --- | --- | --- | --- | --- | --- | --- |
| Parameter (Units) | Day 1 | Day 5 | Day 1 | Day 5 | Day 1 | Day 5 |
|  | N=4 | N=4 | N=4 | N=4 | N=4 | N=3 |
| Molnupiravir (EIDD2801) |  |  |  |  |  |  |
| AUC<sub>0-4</sub> (ng.h/mL) | NC | NC | NC | NC | NC | NC |
| C<sub>max</sub> (ng/mL) | 5.76 (13.3) | NC | 25.8 | 9.14<sup>c</sup><br>(5.97 – 12.3) | 8.43 (58.6) | 7.79 (13.9) |
| T<sub>max</sub><sup>a</sup> (h) | 0.500<br>(0.500-0.500) | NC | 1.00b<br>(1.00-1.00) | 0.500<sup>c</sup><br>(0.500-0.500) | 0.750 (0.500-1.00) | 1.00 (1.00-1.00) |
| NHC (EIDD1931) |  |  |  |  |  |  |
| AUC<sub>0-4</sub> (ng.h/mL) | 3210 (40.5) | 3470 (42.4) | 4610<br>(33.7) | 3880<br>(56.3) | 9240<br>(41.0) | 7880 (39.0) |
| C<sub>max</sub> (ng/mL) | 1490 (29.4) | 1620 (51.0) | 2230<br>(38.2) | 1820<br>(84.6) | 4440<br>(45.2) | 4180 (28.1) |
| T<sub>max</sub><sup>a</sup> (h) | 1.50<br>(1.00-2.00) | 1.00<br>(1.00-2.00) | 1.50<br>(1.00-2.00) | 1.00<br>(1.00-2.00) | 2.00<br>(1.00-2.00) | 2.00<br>(1.00-2.00) |

NA = Not applicable, NC = Not calculable,
AUC<sub>0-4</sub> = Area under the time-concentration curve (0-4h)
T<sub>max</sub> = Time to peak concentration
C<sub>max</sub> – Peak concentration
a Median (min-max) presented,
b n=1 with quantifiable concentrations out of 4 subjects
c n=2 with quantifiable concentrations out of 4 subjects

## DISCUSSION

To study the tolerability and safety of molnupiravir, we enrolled participants who presented within five days of symptoms, and who did not have severe disease, since we judged that the largest public health impact of this antiviral drug would be through deployment in the community for preventing hospitalisation. In untreated SARS-CoV-2 infection, viral load peaks in the first week of illness ^8^ suggesting that early antiviral treatment may influence disease progression and potentially transmission.

We have established the safety and tolerability of molnupiravir in SARS-CoV-2 infected individuals, alongside a conventional phase I dose ranging study in healthy volunteers (NCT04392219). We have shown that a dose of 800mg bd of molnupiravir is safe and well-tolerated in participants with SARS CoV-2 infection; the plasma concentrations attained are within the target range based on scaling from animal models ^2,3^. Adverse effects were commonly reported, affecting 9/12 and 5/6 participants on molnupiravir and controls respectively. All were mild (Grade 1-2) and included flu-like and upper respiratory symptoms, headache, myalgia, diarrhoea and nausea which were also consistent with symptomatic COVID-19 disease.

AGILE utilises complex innovative trial design methodology to accelerate early phase evaluation of novel antiviral agents against SARS CoV-2. Our Bayesian approach was selected to optimise statistical efficiency and to accelerate decision-making. Drug safety is not definitively established during phase I and requires large numbers of individuals dosed in phase III or IV. Rather, the AGILE design allowed us to establish (within an accelerated timescale) that a dose of molnupiravir 800mg bd for 5 days was sufficiently safe to progress into our continuation phase IIa placebo-controlled trial (where safety continues to be monitored). Since full reproductive toxicologic datasets were not available at the time of initiation, our study required stringent precautions to avoid pregnancy in participants or their partners.

To our knowledge this is the first published report describing the use of molnupiravir in SARS-CoV-2 infected individuals. We observed comparable exposures of EIDD1931 to healthy volunteers,^8^and describe an approach for rapidly estimating a dose-toxicity relationship for phase II evaluation. Whether or not molnupiravir will prove effective in treating COVID-19 will be determined in phase II trials which are currently underway, including our own, but the paucity of potent antiviral agents in the COVID-19 pipeline strongly argues for such accelerated approaches to early phase drug development.

## Data Availability

The AGILE Trial Steering Committee will consider all reasonable requests by health-care providers, investigators, and researchers to provide anonymised data to address specific scientific or clinical objectives. The AGILE investigators are committed to reviewing requests from researchers for access to clinical trial protocols, de-identified patient-level clinical trial data, and study-level clinical trial data.

## Acknowledgments

We thank the AGILE Trial Steering Committee (Nicholas Paton (chair), Fred Hayden, Janet Darbyshire, Amy Lucas, Ulrika Lorch), the AGILE Safety Review Committee (Amitava Ganguli, Wendy Painter), our independent Data Monitoring and Ethics Committee (Andrew Freedman (chair), Richard Knight, Richard Peck, Stevan Julious), and the AGILE Scientific Advisory Board (Annalisa Jenkins (chair), Stevan Emmett, Sanjay Bhagani, Sheuli Porkess, Larry Zeitlin, Gary Kobinger).

We thank the following for trial support: Kelly Byrne, Karen Martin, Kelly Cozens, Sam Wilding, Michelle Light, Cleo Pike, Nadia Kontogianni, Rachel Byrne, Ana I Cubas-Atienzar, Thomas Edwards, Jayne Jones.

We acknowledge National Institute for Health Research infrastructure funding for the Liverpool Clinical Research Facility and the Southampton Clinical Trials Unit. We thank Dr John Powers, Leidos Biomedical and the National Institute for Allergy and Infectious Diseases (NIAID), National Institutes for Health for supplying the FLU-PRO Questionnaire. TJ was supported by the NIHR Cambridge Biomedical Research Centre (BRC-1215-20014) and by UK Medical Research Council (grant number: MC_UU_00002/14). The views expressed are those of the author(s) and not necessarily those of the NIHR or the Department of Health and Social Care.

## Contributors Statement

SK, GG, TJ, SE, RF, PM contributed to study design. SK, GG, TJ, SE, GS, KT, PM, HP contributed to data analysis and interpretation. RF led clinical conduct as the principal investigator of the clinical site. TF, LW, RL, MB participated in clinical assessment and data collection. KF participated in the management of pharmacovigilance. SC, EW, MR, LJ contributed to the digital data collection and data management of the trial. MR led set up of the randomisation system. WG, TF, VS, EA, KB, CH contributed to study bioanalysis. AC, ND, EM, OTH, SY, HR, JC, RL, CW, JB, AO, MJ, DGL contributed to study management and execution. KM contributed to monitoring activities. WP, WH contributed preclinical and safety data on molnupiravir. SK, GG, MJ, AO, DGL were involved in primary manuscript writing. All authors contributed to the final version of the manuscript.

## Supplementary Information Titles

S1: Model-based dose-finding design

S2: Clinical endpoints day 15 and day 29 (Evaluable population)

